# Structural variants linked to Alzheimer’s Disease and other common age-related clinical and neuropathologic traits

**DOI:** 10.1101/2024.08.12.24311887

**Authors:** Ricardo A Vialle, Katia de Paiva Lopes, Yan Li, Bernard Ng, Julie A Schneider, Aron S Buchman, Yanling Wang, Jose M Farfel, Lisa L Barnes, Aliza P Wingo, Thomas S Wingo, Nicholas T Seyfried, Philip L De Jager, Chris Gaiteri, Shinya Tasaki, David A Bennett

## Abstract

Advances have led to a greater understanding of the genetics of Alzheimer’s Disease (AD). However, the gap between the predicted and observed genetic heritability estimates when using single nucleotide polymorphisms (SNPs) and small indel data remains. Large genomic rearrangements, known as structural variants (SVs), have the potential to account for this missing genetic heritability. By leveraging data from two ongoing cohort studies of aging and dementia, the Religious Orders Study and Rush Memory and Aging Project (ROS/MAP), we performed genome-wide association analysis testing around 20,000 common SVs from 1,088 participants with whole genome sequencing (WGS) data. A range of Alzheimer’s Disease and Related Disorders (AD/ADRD) clinical and pathologic traits were examined. Given the limited sample size, no genome-wide significant association was found, but we mapped SVs across 81 AD risk loci and discovered 22 SVs in linkage disequilibrium (LD) with GWAS lead variants and directly associated with AD/ADRD phenotypes (nominal *P* < 0.05). The strongest association was a deletion of an *Alu* element in the 3’UTR of the *TMEM106B* gene. This SV was in high LD with the respective AD GWAS locus and was associated with multiple AD/ADRD phenotypes, including tangle density, TDP-43, and cognitive resilience. The deletion of this element was also linked to lower TMEM106B protein abundance. We also found a 22 kb deletion associated with depression in ROSMAP and bearing similar association patterns as AD GWAS SNPs at the *IQCK* locus. In addition, genome-wide scans allowed the identification of 7 SVs, with no LD with SNPs and nominally associated with AD/ADRD traits. This result suggests potentially new ADRD risk loci not discoverable using SNP data. Among these findings, we highlight a 5.6 kb duplication of coding regions of the gene *C1orf186* at chromosome 1 associated with indices of cognitive impairment, decline, and resilience. While further replication in independent datasets is needed to validate these findings, our results support the potential roles of common structural variations in the pathogenesis of AD/ADRD.

## BACKGROUND

Alzheimer’s disease (AD) is a complex disorder heavily influenced by genetics. While twin studies have estimated the broad-sense heritability of AD between 60-80% (Gatz et al. 2006), SNP-based heritability varies across different studies, ranging from 5-30% (Escott-Price and Hardy 2022; Wang et al. 2021). Epistatic interactions, unbalanced age-matched controls, and the large proportion of "proxy” cases are among the reasons that could explain such a gap between the expected and observed heritability (Escott-Price and Hardy 2022). Large genomic rearrangements, known as structural variants (SVs), can also account for some missing genetic heritability. This class of variants, often not discoverable using genotyping assays, is recently garnering attention with the popularization of whole-genome sequencing data which allows detection of such variants at a large scale with considerable confidence (Collins et al. 2020; Sudmant et al. 2015).

Although a few SVs have already been linked to AD, with the most well-known being rare duplications of the *APP* gene causally linked to the early onset form of the disease (Blom et al. 2008; Hooli et al. 2012; Kasuga et al. 2009; Rovelet-Lecrux et al. 2006; Sleegers et al. 2006), our knowledge of the impact of common SVs in late-onset AD (LOAD) is mostly limited to a few copy number variants (CNVs), which are usually detected using SNP arrays or PCR assays and identified in small sample-size studies that have weak replication (Wang et al. 2022). For other neurodegenerative diseases, the role of SVs is usually clearer. For example, in frontotemporal dementia (FTD), repeat expansions in *C9orf72* and the *MAPT* locus inversion are linked to disease risk (Baker et al. 1999; DeJesus-Hernandez et al. 2011). The same inversion is also a major genetic risk factor for progressive supranuclear palsy (PSP) (Baker et al. 1999), and it is associated with Parkinson’s disease (PD) (Zabetian et al. 2007). Additionally, a recent study found a common deletion in the gene *TPCN1* associated with Lewy body dementia (LBD) in a locus also associated with AD GWAS (Bellenguez et al. 2022; Kaivola et al. 2023). These findings highlight the fact that mapping SVs across multiple diseases and related clinical and neuropathological traits can be beneficial to understanding their role in complex diseases.

To investigate the role of SVs in AD/ADRD traits, we leveraged data from two ongoing cohort studies of aging and dementia: the Religious Orders Study and the Rush Memory and Aging Project (ROS/MAP). These cohorts benefit from having a comprehensive set of phenotypes measured from the same set of individuals. We previously reported that SVs have an impact on many molecular phenotypes in the human brain, including over 300 SVs that were also in LD with GWAS traits (Vialle et al. 2022). Here we extend our findings in many ways by effectively linking SVs to an extensive list of AD/ADRD phenotypes measured for the same individuals. Over 20 thousand common SVs mapped in 1,088 samples with whole genome sequencing (WGS) data were tested for association with multiple indices for AD/ADRD, including clinical diagnoses of AD dementia and mild cognitive impairment (MCI); multiple measurements of cognition (e.g., cognitive decline and resilience); diagnosis of major depressive disorder (MDD) and depressive symptoms; indices of motor function; neuropathological evaluations of β-amyloid load, neurofibrillary tangles density, Lewy Bodies and TDP-43; and indices of cerebrovascular diseases (CVD) such as cerebral amyloid angiopathy (CAA), gross and microinfarcts, atherosclerosis, and arteriolosclerosis.

## RESULTS

### Characteristics of the samples

Genotyped structural variant calls were obtained on 1,088 non-Latino white subjects from the ROS/MAP cohort studies (David A. Bennett et al. 2018). In both studies, participants underwent annual clinical evaluations and donated their brains at death. The mean (SD) age at enrollment and age at death across the participants used in this study was 80.9 (6.8) and 89.0 (6.4) years, respectively, with an average (SD) follow-up period of 7.2 (4.9) years. Of all participants, 43.7% had a diagnosis of Alzheimer’s dementia at death, and nearly two-thirds had pathologic AD confirmed post-mortem. A decline in cognition was observed, with the Mini-Mental State Examination (MMSE) score decreasing from 28 (IQR 26-29) at baseline to 25 (IQR 15-28) proximate to death. TDP-43 pathology extending beyond the amygdala was observed in just over a third of brains, and 13% presented Lewy bodies in nigra and/or cortex regions. Cerebrovascular diseases, including macroscopic infarcts and microinfarcts, were observed in more than a third and a quarter, respectively, and moderate to severe amyloid angiopathy, atherosclerosis, and arteriolosclerosis were observed in about a third of the brains. Detailed characteristics of each sample are presented in **Supplementary Table 1**.

### Genome-wide SV association scans with clinical and neuropathologic phenotypes

Genome-wide association scans of structural variants were performed on a range of clinical and neuropathologic phenotypes covering multiple clinical and pathologic variables related to aging and dementia. Twenty four phenotypes were analyzed, including clinical diagnosis of AD, MCI, and MDD; depressive symptomatology; measurements of cognitive function, motor function, frailty and parkinsonian scores; neuropathologies, such as β-amyloid, tangles, TDP-43, Lewy Bodies, and multiple cerebrovascular diseases indices. SV discovering and genotyping pipelines were obtained from combined ROS/MAP samples as previously described (Vialle et al. 2022). SVs were classified into different classes of variation, including deletions (DEL), insertions (INS), duplications (DUP), inversions (INV), complex rearrangements (CPX), and three classes of mobile element insertions (MEI), *Alu*, SVA, and LINE1. A total of 72,348 SVs were initially mapped. For the association analysis, variants with Hardy-Weinberg equilibrium (HWE) *P*-value lower than 10^-6^ and minor allele frequency (MAF) lower than 1% were removed. Single variant association tests were applied across 16 quantitative and eight binary traits (**Supplementary Table 2**) for about 20 thousand SVs with minor allele count (MAC) greater than 10 in each cohort. Scans were performed using the tool SAIGEgds (Scalable and Accurate Implementation of GEneralized mixed model) (Zheng and Davis 2021; Zhou et al. 2018). All tests were controlled by age at death, sex, years of education, five genetic principal components, and the genetic correlation matrix (modeled as a random effect). A meta-analysis was performed combining results from both ROS and MAP cohorts. Genomic inflation (λ) ranged from 0.59 to 1.08, indicating a reasonable control of population stratification and relatedness (**Table 1**). None SVs reached genome-wide significance (*P* < 5 x 10^-8^) in any of the samples and phenotypes tested at the current sample size. Complete summary stats for each phenotype are provided in **Additional Data** (see Data availability).

**Table 1.** Genomic inflation of genome wide association tests in each cohort and meta-analysis.

| Phenotype | ROS ($\lambda$) | MAP ($\lambda$) | Meta-Analysis (ROS/MAP) ($\lambda$) |
| --- | --- | --- | --- |
| *Cognition* |  |  |  |
| Alzheimer's Dementia | 1.01 | 1.01 | 0.99 |
| Mild cognitive impairment | 1.01 | 0.99 | 0.98 |
| Global cognitive function | 1.00 | 0.73 | 0.89 |
| Cognitive decline | 1.02 | 1.02 | 0.97 |
| Cognitive resilience | 1.02 | 1.01 | 1.03 |
| *Depression* |  |  |  |
| Depressive symptoms (CES-D) | 0.67 | 1.03 | 1.01 |
| Major Depressive Disorder | 1.08 | 1.02 | 1.03 |
| *Motor* |  |  |  |
| Frailty | 0.59 | 0.99 | 1.02 |
| Motor functions | 0.90 | 1.01 | 0.94 |
| Global parkinsonian score | 0.91 | 0.85 | 1.00 |
| *Pathology* |  |  |  |
| NIA-Reagan diagnosis of AD | 1.01 | 1.03 | 0.99 |
| Global AD pathology | 0.99 | 1.00 | 0.99 |
| Beta-Amyloid | 1.01 | 1.00 | 1.01 |
| Tangle density | 1.03 | 0.99 | 0.99 |
| Diffuse plaques | 1.01 | 0.91 | 0.99 |
| Neuritic plaques | 1.01 | 1.02 | 1.00 |
| Neurofibrillary tangle | 1.01 | 0.86 | 0.92 |
| TDP-43 | 1.01 | 0.99 | 1.01 |
| Lewy Body disease | 0.99 | 1.00 | 1.00 |
| *Vascular Pathology* |  |  |  |
| Arteriolosclerosis | 1.03 | 0.96 | 0.99 |
| Cerebral Atherosclerosis | 1.03 | 0.99 | 1.00 |
| Cerebral amyloid angiopathy | 1.00 | 0.99 | 1.00 |
| Cerebral Infarctions (Gross) | 1.01 | 1.01 | 1.00 |
| Cerebral Infarctions (Micro) | 1.00 | 1.00 | 1.00 |

### SVs in AD GWAS *loci*

To investigate the impact of SVs in known AD GWAS *loci*, we examined 81 genome-wide significant loci collectively identified in six previous studies that did not examine SVs (Andrews et al. 2023). We first mapped the presence of SVs (discovered in ROS/MAP) in each locus. On average, 10 SVs were identified by locus across all 81 loci (**Supplementary Figure S1**). As expected, complex genomic regions, such as the *HLA* locus, harbored a considerably higher number of SVs (75), followed by the *ABCA7* (47), the *TMEM121* (35), and *IDUA* (32) loci. 36 SVs were in LD with the lead variant in 10 of the 81 loci, with R^2^ ranging from 0.217 to 0.964. Among these, 22 SVs were nominally associated (*P* < 0.05) with at least one of the 24 AD/ADRD phenotypes tested (**Figure 1** and **Supplementary Table 3**).

**Figure 1.**
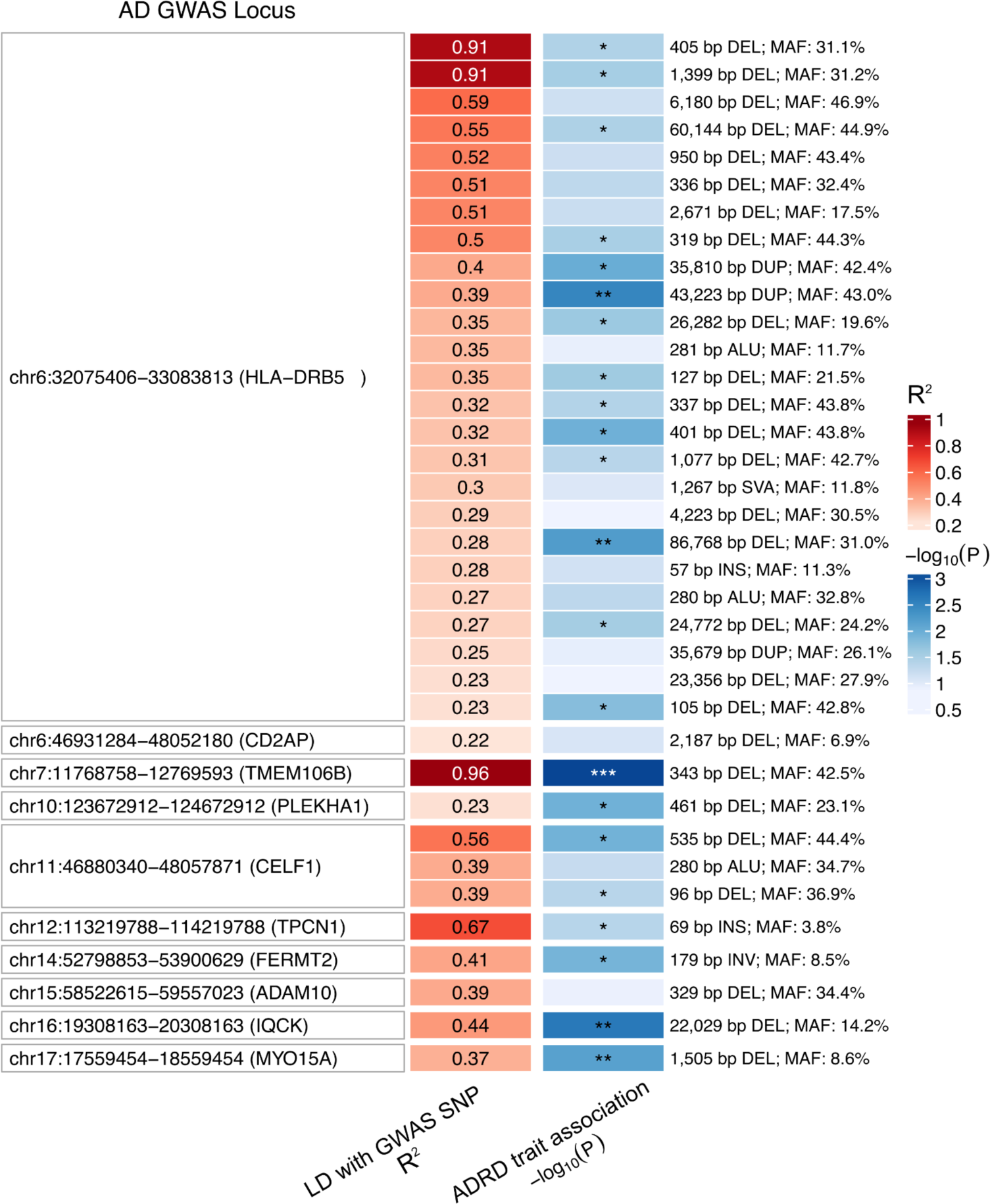
Structural variants in LD with AD GWAS variants. Heatmap shows 36 SVs in LD (R^2^ > 0.2) with 10 AD GWAS loci lead SNPs. The best association of each SV with ROS/MAP AD/ADRD phenotypes is highlighted in the blue colored column (shown as −log_10_(nominal *P*); \**P* ≤ 0.05, \*\**P* ≤ 0.01, \*\*\**P* ≤ 0.001).

The SV with the strongest results (*P* = 7.72 x 10^-4^) was a 343 bp deletion (**Figure 2C**), which deletes an *Alu* element at the 3’UTR of *TMEM106B* (Rodney et al. 2024), and was in high LD with the lead variant at the *TMEM106B* locus (rs5011436; R^2^ = 0.96) (**Figure 2A**). In ROS/MAP participants, this SV was associated with multiple AD/ADRD phenotypes, including tangle density, cognitive resilience, TDP-43, and others, illustrating the pleiotropy among SVs (**Figure 2B**). We further collected information on SV-xQTL and found that the deletion was also associated with lower protein abundance of *TMEM106B* (**Figure 2D**).

**Figure 2.**
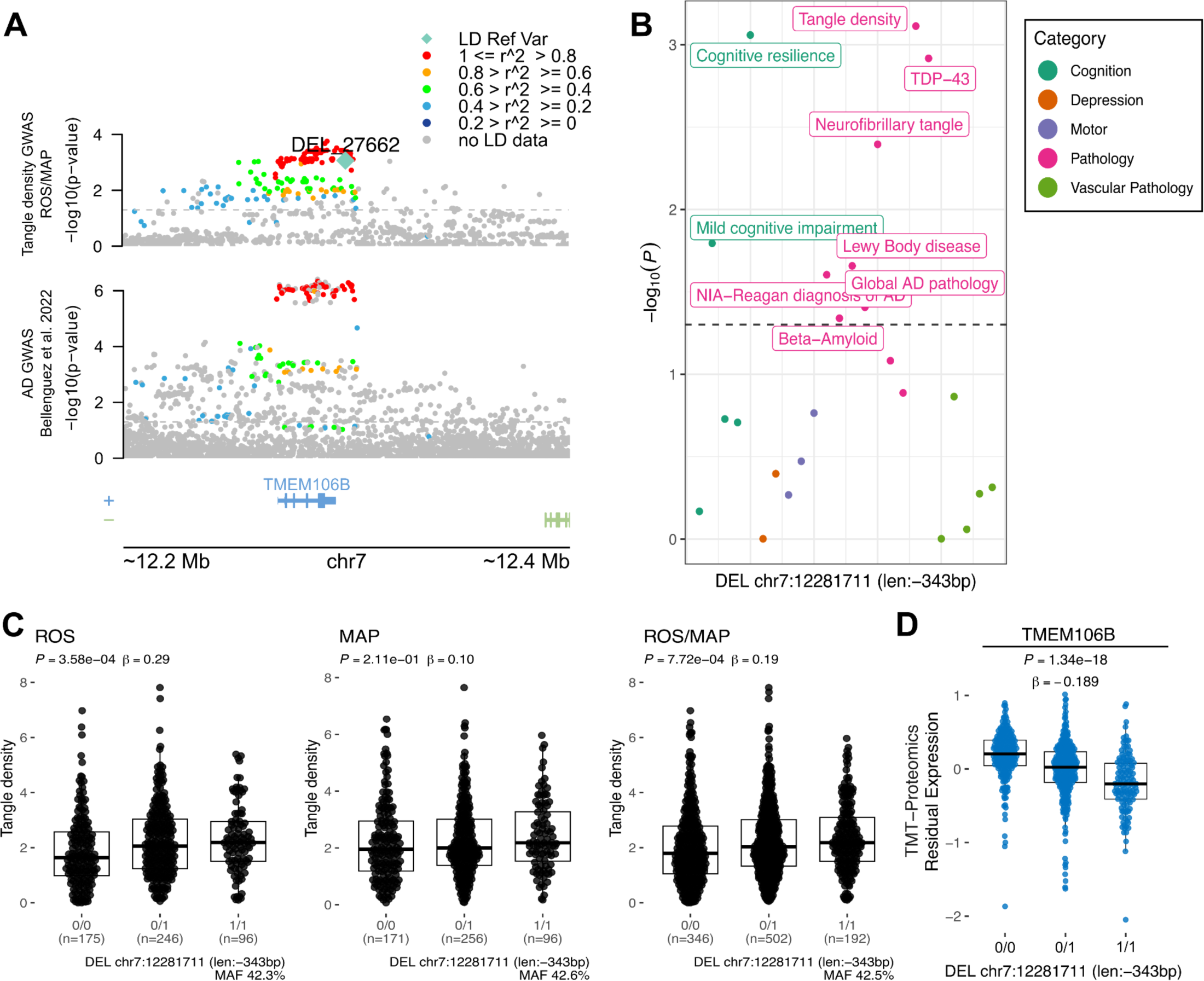
343 bp deletion associated with tangle density at *TMEM106B* locus. **A**, locus zoom plot for the *TMEM106B* locus (chr7:11,768,758-12,769,593) showing a 200 Kbp window around a 343 bp deletion with high LD with the lead SNP rs5011436. On the top, the y-axis shows the nominal *P* values (as −log_10_) for the association tests with tangle density in ROS/MAP participants. On the bottom, the y-axis shows the AD GWAS results from Bellenguez et al. (2022). The SV is plotted in a diamond shape, while SNPs are plotted in circles. Points are colored by the LD (R^2^) to the SV. The dashed line represents nominal *P* = 0.05. **B**, shows the nominal *P* values (as −log_10_) for the association of the SV with all AD/ADRD traits tested. Dots are colored by phenotype category. The dashed line represents nominal *P* = 0.05. **C**, Boxplots showing the tangle density measures by the deletion alleles (ROS, MAP, ROS/MAP). **D**, Boxplot shows the SV-pQTL between the deletion and protein expression levels of *TMEM106B* measured from DLPFC brain tissues of 663 ROS/MAP participants.

Another four associations were found with a relaxed *P* < 0.01. That included a 22,029 bp long deletion (**Figure 3A**) associated with diagnosis of MDD in ROS/MAP (*P* = 0.0025) and in LD (R^2^ = 0.44) with the lead variant in the *IQCK* locus (chr16:19,308,163-20,308,163)(**Figure 3B**). Two SVs at the HLA locus, an 86,768 bp deletion (R^2^ = 0.27) and a 43,223 bp duplication (R^2^ = 0.39) associated with cognitive resilience (*P* = 0.002) and MDD (*P* = 0.003), respectively. And one 1,505 bp deletion at the MYO15A locus (R^2^ = 0.37), associated with TDP-43 (*P* = 0.007). Although the AD association at the *IQCK* locus was identified only in one paper (Kunkle et al. 2019), there are still nominally significant associations at more powered AD GWAS (**Figure 3C**). In ROS/MAP, apart from depression status, cognitive resilience and cognitive decline also reached a nominal significant threshold (**Figure 3D**). Depression is a well-known risk factor for AD, associated with an increased likelihood of developing dementia (Bellou et al. 2017; Dafsari and Jessen 2020; Harerimana et al. 2022; Holmquist, Nordström, and Nordström 2020), and this association could represent a genetic link between depression and AD. The SVs at the HLA locus overlap the genes *HLA-DRB1*, *HLA-DRB5*, and *HLA-DRB6*, characterizing the class II sub-region haplotypes. However, their association and LD patterns are overly complex to disentangle the causal variants from the haplotypes.

**Figure 3.**
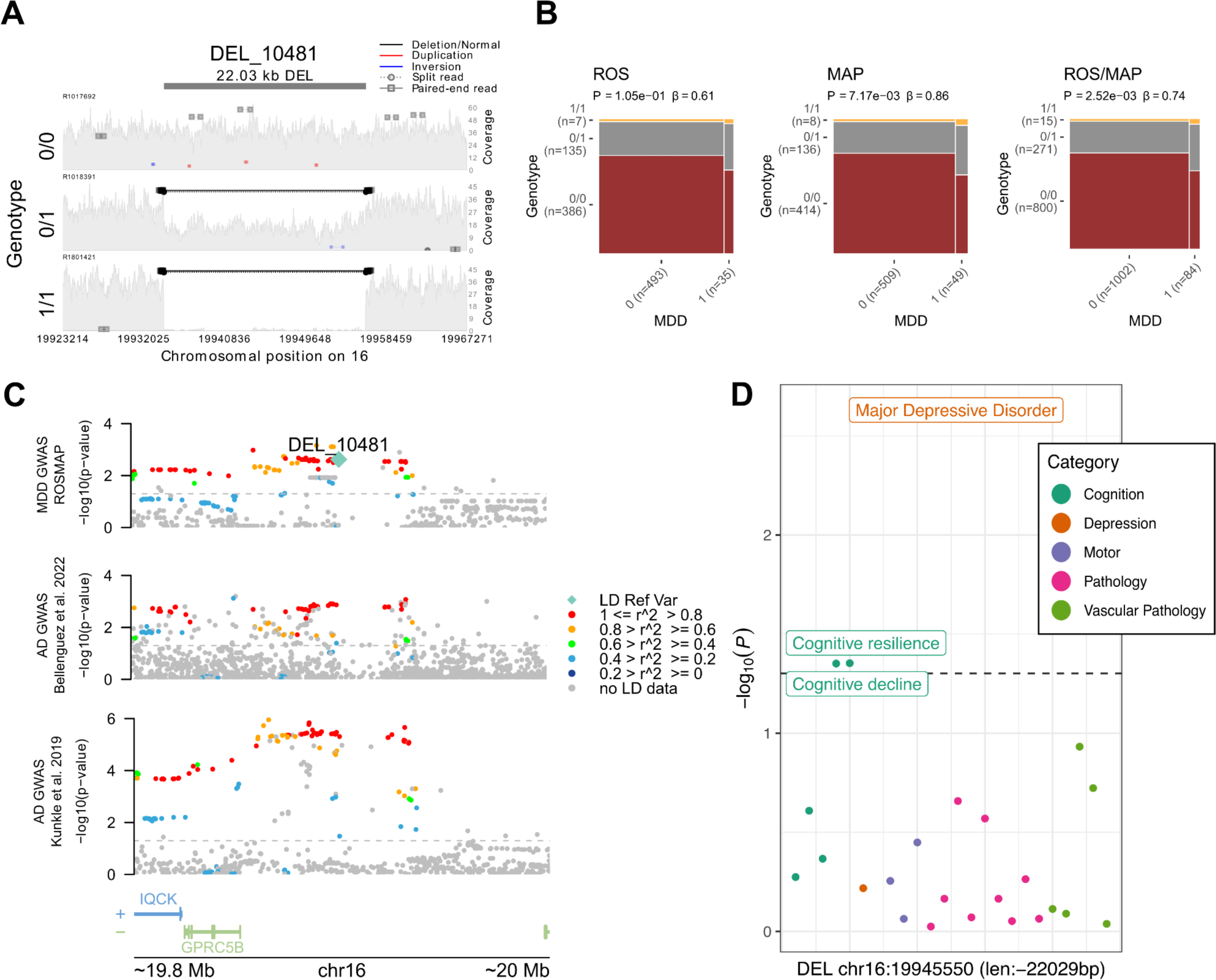
22 Kbp deletion associated with major depressive disorder at *IQCK* locus. **A**, mapping of sequencing reads at the locus from three representative individuals corresponding to the possible genotypes: deletion not present (0/0), heterozygous deletion (0/1), and homozygous deletion (1/1). **B**, mosaic plots showing the proportion of individuals with MDD and respective deletion alleles for ROS, MAP, and ROS/MAP. **C**, locus zoom plot for the AD GWAS *IQCK* locus (chr16:19,308,163-20,308,163) showing a 200 Kbp window around the deletion in LD with the lead SNP rs7185636. The scatter plot on the top shows the nominal *P* values (as −log_10_) for the association tests with MDD status in ROS/MAP participants. The scatter plots in the middle and bottom show AD GWAS results from Bellenguez et al. (2022) and Kunkle et al. (2019), respectively. The 22 Kbp deletion is plotted in a diamond shape, while SNPs are plotted in circles. Points are colored by the LD (R^2^) to the SV, as measured in ROS/MAP. The dashed line represents nominal *P* = 0.05. **D**, shows the nominal *P* values (as −log_10_) for the association of the 22 Kbp deletion with all AD/ADRD traits tested. Dots are colored by phenotype category. The dashed line represents nominal *P* = 0.05.

### Genome-wide association SV reveals potentially novel ADRD risk loci

As an exploratory analysis, we performed a meta-analysis combining association results from 529 ROS participants and 559 individuals from MAP. We were able to map suggestive associations that could represent potential novel risk loci by focusing on SVs with no LD with SNPs and a relaxed *P*-value threshold of *P* < 5 x 10^-3^ with at least one ADRD phenotype. To verify the robustness of the variant call and avoid false positives, we selected only SVs also mapped in gnomAD (with 80% reciprocal overlap). That resulted in 9 SV-phenotype associations comprising 7 SVs (**Table 2**; **Figure 4**), four mapped to intergenic regions, two intronic, and one with overlaps to coding regions. SV lengths ranged from 69 bp to 5.6 kb, with MAFs from 5.2% to 42.4%.

**Figure 4.**
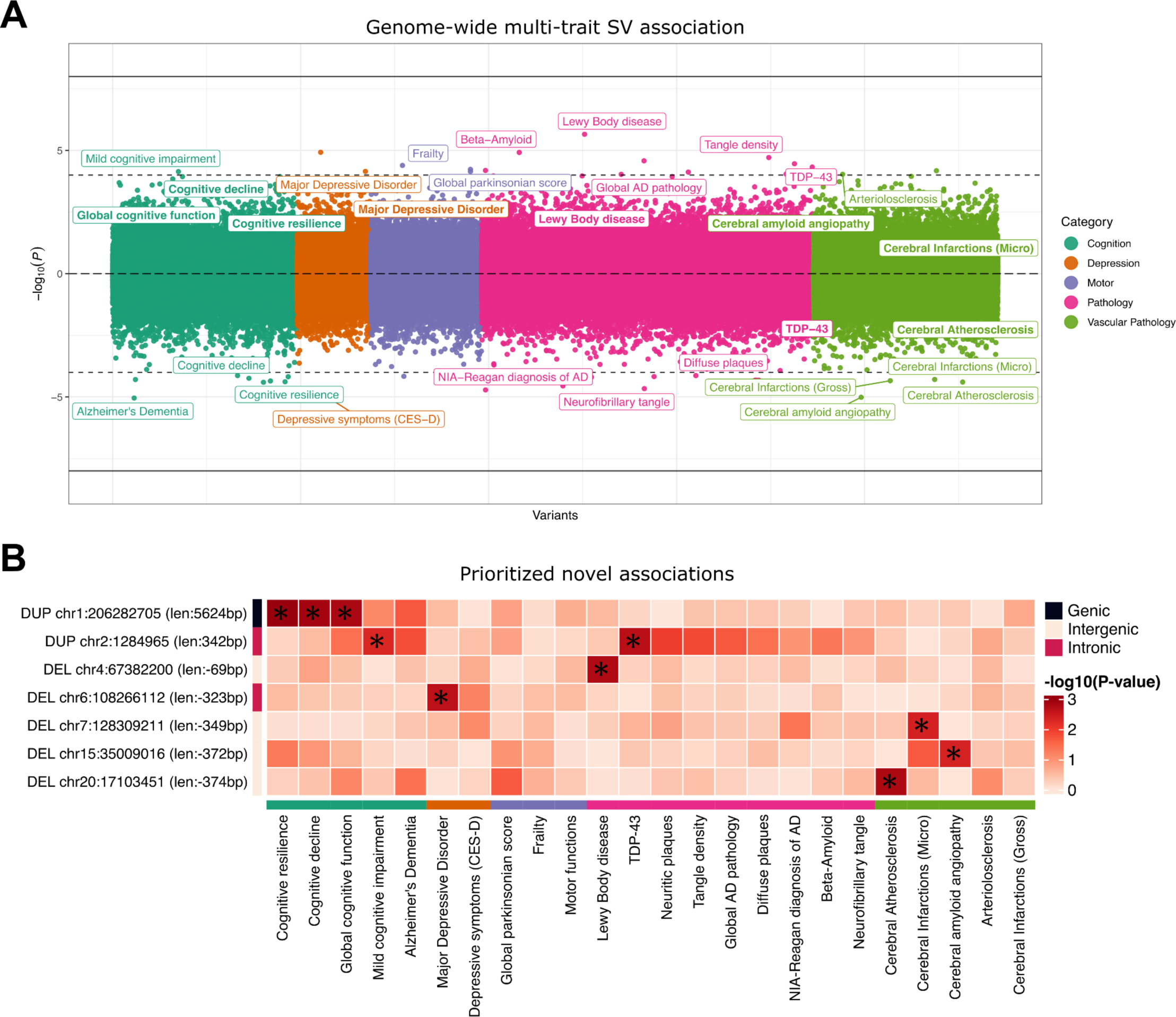
Chicago plot showing the direction of associations with results of the SV genome-wide scan in 1,088 ROS/MAP participants across 24 AD/ADRD phenotypes. **A**, Phenotypes are colored by common related categories. Solid-line represents the genome-wide significance of *P* < 5 x 10^-8^. The dashed line represents a suggestive threshold of *P* < 10^-4^. Phenotype labels are shown only for associations below the suggestive threshold. Prioritized associations (*P*_META_ < 5 x 10^-5^ and no LD with SNPs) are highlighted in bold. **B**, List of 7 prioritized SV-trait associations under the conditions of *P*_META_ < 5 x 10^-3^ for at least one ADRD phenotype, no LD with SNPs in their respective loci, and supporting evidence of the SV call in gnomAD. Bonferroni adjusted *P*-values are displayed in the heatmap. Asterisks indicate adj. *P* < 0.05.

**Table 2.** SVs associated with AD/ADRD traits with no LD with SNPs.

| Phenotype | SV | MAF | ROS |  | MAP |  | Meta-Analysis<br>(ROS/MAP) |  | Gene |
| --- | --- | --- | --- | --- | --- | --- | --- | --- | --- |
|  |  |  | Beta (SE) | P | Beta (SE) | P | Beta (SE) | P |  |
| Cognitive resilience | DUP_267<br>chr1:206282705<br>(len:5,624bp) | 34.3% | 0.01 (0.01) | $1.27 \times 10^{-1}$ | 0.02 (0.01) | $2.41 \times 10^{-3}$ | 0.01 (0.00) | $1.02 \times 10^{-3}$ | *C1orf186* |
| Cognitive decline | DUP_267<br>chr1:206282705<br>(len:5,624bp) | 34.3% | 0.02 (0.01) | $3.90 \times 10^{-2}$ | 0.02 (0.01) | $1.11 \times 10^{-2}$ | 0.02 (0.01) | $1.28 \times 10^{-3}$ | *C1orf186* |
| Global cognition | DUP_267<br>chr1:206282705<br>(len:5,624bp) | 34.3% | 0.26 (0.12) | $2.96 \times 10^{-2}$ | 0.29 (0.12) | $1.71 \times 10^{-2}$ | 0.27 (0.08) | $1.51 \times 10^{-3}$ | *C1orf186* |
| Cerebral Atherosclerosis | DEL_17353<br>chr20:17103451<br>(len:-374bp) | 8.5% | -0.20 (0.09) | $2.41 \times 10^{-2}$ | -0.22 (0.10) | $2.20 \times 10^{-2}$ | -0.21 (0.07) | $1.58 \times 10^{-3}$ | *RNU6-27P* |
| Lewy Body disease | DEL_21899<br>chr4:67382200<br>(len:-69bp) | 28.3% | 0.49 (0.22) | $2.44 \times 10^{-2}$ | 0.46 (0.20) | $2.42 \times 10^{-2}$ | 0.46 (0.15) | $1.73 \times 10^{-3}$ | *RPS23P3* |
| TDP-43 | DUP_2394<br>chr2:1284965<br>(len:342bp) | 42.4% | -0.70 (0.27) | $8.19 \times 10^{-3}$ | -0.41 (0.22) | $6.40 \times 10^{-2}$ | -0.53 (0.17) | $2.19 \times 10^{-3}$ | *SNTG2* |
| MDD | DEL_26626<br>chr6:108266112<br>(len:-323bp) | 28.2% | 0.78 (0.36) | $2.85 \times 10^{-2}$ | 0.71 (0.32) | $2.81 \times 10^{-2}$ | 0.76 (0.24) | $2.31 \times 10^{-3}$ | *SEC63* |
| Micro-chronic cerebral infarctions | DEL_28995<br>chr7:128309211<br>(len:-349bp) | 8.6% | -0.10 (0.25) | $6.98 \times 10^{-1}$ | 0.95 (0.26) | $2.87 \times 10^{-4}$ | 0.40 (0.18) | $4.29 \times 10^{-3}$ | *RP11-274B21.5* |
| Cerebral amyloid angiopathy | DEL_9454<br>chr15:35009016<br>(len:-372bp) | 5.2% | 0.36 (0.14) | $8.63 \times 10^{-3}$ | 0.19 (0.13) | $1.27 \times 10^{-1}$ | 0.26 (0.09) | $4.40 \times 10^{-3}$ | *GJD2* |

The most promising finding was a 5.6 kb duplication at the 1q31.1 locus, which completely overlaps a non-canonical exon and parts of the 5’UTR of the gene *C1orf186* (which codes for the protein RHEX)(**Figure 5A-B**). This duplication was associated with better measures of cognitive resilience (*P*_ROS_ = 1.27 × 10^−1^, *P*_MAP_ = 2.41 x 10^-3^, *P*_META_ = 1.02 x 10^-3^) (**Figure 5C**), and also slopes of cognitive decline and global cognition proximal to death (**Figure 5D** and **Table 2**). MAF in both studies was around 34%, with no homozygous carriers with measures of resilience available. Given that *C1orf186* is not expressed in brain tissues, no QTL mapping was measured between the duplication with the expression of *C1orf186* in ROS/MAP brains. Other suggestive findings included two SVs overlapped intronic regions, a 342 bp duplication in an intronic region of the gene *SNTG2* at chr2 associated with TDP-43 and several other AD neuropathologies, and a 323 bp deletion in an intronic region of the gene *SEC63* at chr6 associated with MDD. Finally, four other SVs were mapped to intergenic regions, a 374 bp deletion at chr20 associated with cerebral atherosclerosis; a 69 bp deletion at chr4 associated with the presence of Lewy bodies; a 349 bp deletion at chromosome 7 overlapping associated with micro-chronic cerebral infarctions; and a 372 bp deletion at chr15 associated with cerebral amyloid angiopathy.

**Figure 5.**
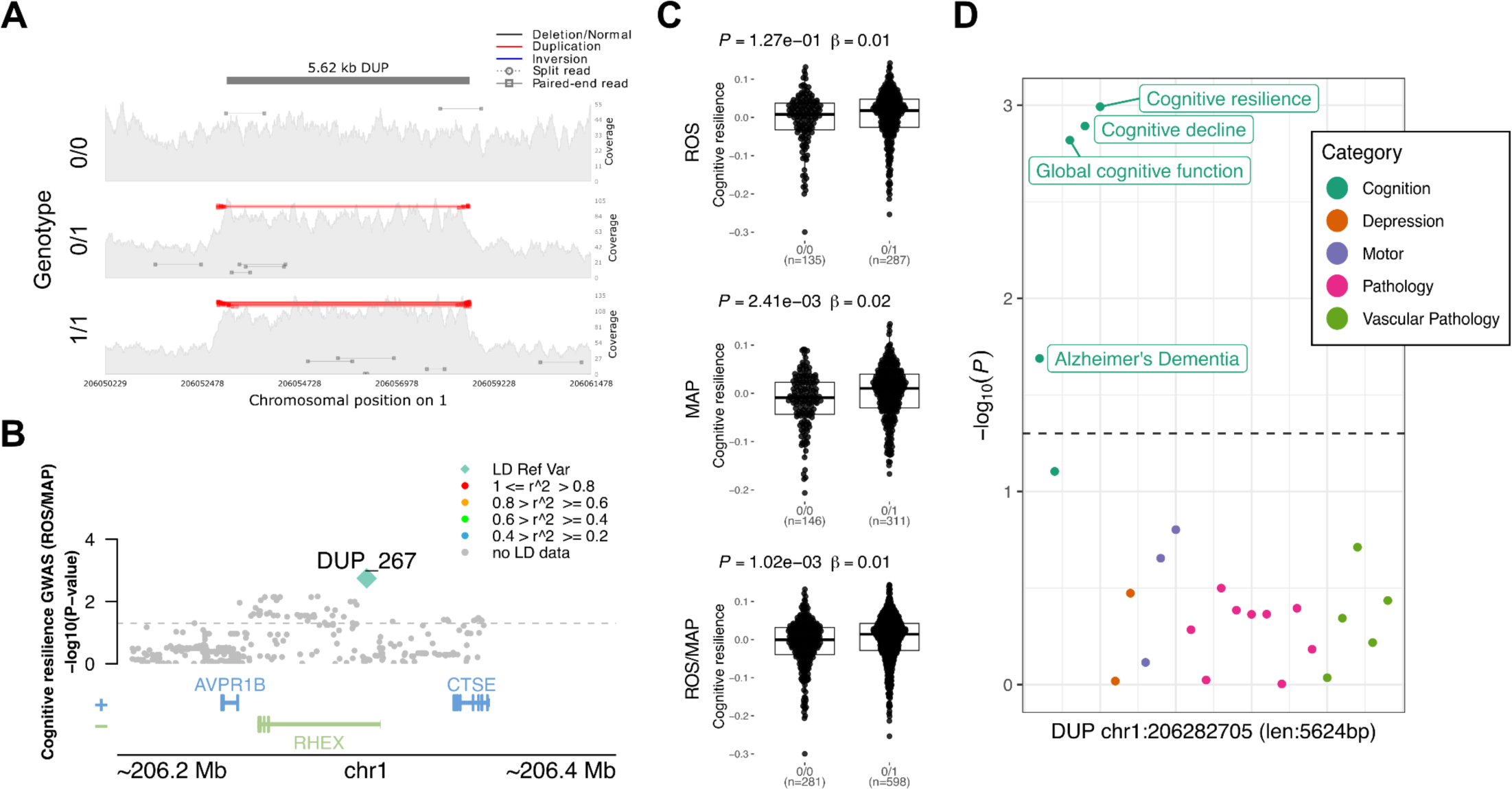
5.6 Kbp duplication at the 1q31.1 locus associated with cognitive resilience. **A**, mapping of sequencing reads at the locus from three representative individuals corresponding to the possible genotypes: duplication not present (0/0), heterozygous allele (0/1), and homozygous allele (1/1). **B**, locus zoom plot showing 200 Kbp window around the SV. The y-axis shows the nominal *P* values (as −log_10_) for the association tests with cognition in ROS/MAP participants. The SV is plotted in a diamond shape, while SNPs are plotted in circles. Points are colored by the LD (R^2^) to the SV. **C**, Boxplots showing the cognitive resilience measures by the duplication alleles (ROS, MAP, ROS/MAP). **D**, shows the nominal *P* values (as −log_10_) for the association of the SV with all other AD/ADRD traits tested. Dots are colored by phenotype category. The dashed line represents nominal P = 0.05.

## DISCUSSION

GWAS have enabled the identification of dozens of common SNVs and small indels contributing to AD/ADRD traits (Bellenguez et al. 2022; Chia et al. 2021; Farrell et al. 2022; Howard et al. 2019; Nalls et al. 2019; Sherva et al. 2023; Zhang et al. 2022; Yan et al. 2021). The contribution of SVs, by contrast, lags. Previous studies were restricted mainly to either CNVs, as in autism (Marshall et al. 2008); rare SVs, as in schizophrenia(Halvorsen et al. 2020; Sebat, Levy, and McCarthy 2009; Walsh et al. 2008); or repeat expansions, as in amyotrophic lateral sclerosis (ALS), FTD, and Huntington’s disease. Here, by leveraging the deep ROS/MAP phenotypic data, we performed for the first time a comprehensive analysis of the role of SVs in AD/ADRD traits by linking common SVs alleles to multiple clinical and neuropathological traits.

We investigated the impact of SVs in known AD GWAS loci. By mapping the presence of SVs in each of the 81 loci, we identified 26 SVs in moderate to high LD with GWAS lead SNPs and directly associated (nominal *P* < 0.05) with AD/ADRD phenotypes in ROS/MAP. The most significant SV was a 343 bp deletion at the 3’UTR of *TMEM106B*, associated with multiple phenotypes, including cognitive resilience, tangle density, and TDP-43, and in high LD with the lead variant at the locus. This SV has already been reported as being the likely causal variant at the locus, with reported risks for frontotemporal lobar dementia with TDP-43 inclusions (FTLD-TDP) (Chemparathy et al. 2023) and neurodegeneration (Salazar et al. 2023). The proposed mechanism involved is via an aging-related negative feedback loop mediated by the presence of the *AluYb8* that dysregulates TDP-43 due to increasing demethylation of *TMEM160B* (Salazar et al. 2023). *TMEM106B* variants have also been reported to affect cell fraction in brain tissues from ROS/MAP, specifically for a subpopulation of excitatory neurons (Fujita et al. 2024; Green et al. 2023).

Our integrated results identified suggestive associations with the potential to be novel ADRD risk loci. We highlight seven SVs with sizes ranging from 69 bp to 5,624 bp. Not surprisingly, most of these were found in non-coding regions, two intronic and four intergenic. Interestingly, a large duplication overlapped the UTR and the coding exonic regions of the gene *C1orf186*. This duplication was consistently associated with cognition-related phenotypes, including cognitive decline and resilience, and seemed to provide a protective effect to AD. The *C1orf186* gene encodes for the RHEX protein, which is involved in the erythropoietin-mediated signaling pathway and positive regulation of erythrocyte differentiation (Verma et al. 2014), and although it is not expressed in brain tissues, that does not exclude the possibility of the duplication affecting expression in other tissues.

Recently, a study analyzed associations of common SVs with PD (Billingsley et al. 2023), finding three deletions reaching genome-wide significance. Another study focused on non-Alzheimer’s dementia, including almost 6 thousand individuals, identified a novel deletion associated with LBD and replicated known loci associated with FTD and ALS (Kaivola et al. 2023). Unfortunately, our study could not test these findings due to differences in variant calling methods. Still, besides those highlighted results, many other potentially interesting findings can still be explored from the genome-wide scans, including SVs in moderate to high LD with SNVs in their loci, where SVs having equal or higher significance with the traits tested might suggest potential causal effects. Although the association tests were restricted to the ROS/MAP samples due to the specificity of phenotypic data, the meta-analysis findings still provide a better selection of candidate variants with consistent effects between two cohort design studies.

While our results represent a step forward in understanding the effects of common genetic variation in AD/ADRD traits, important limitations must be noted: 1) the power for association discovery is constrained by the current sample size; 2) the replication of associations in independent samples is limited to available AD-related phenotypes and might not capture the same nuances from ROS/MAP; 3) SV calling is restricted to deletions, insertions, inversions, and duplication and is still prone to falsely discovered variants and low sensitivity (especially for insertions); 4) the suggestive associations do not represent suggestive causal effects on the traits, especially when LD is present, which would require a more precise fine-mapping analysis; 5) analyses were restricted to germline common autosomal structural variation; 6) since the individuals in this study have a European genetic background, these associations might not transfer to ancestrally diverse population-based data. These limitations could be overcome with additional sample size and deeper sequencing data (e.g., long-reads). Most importantly, these results should be interpreted as suggestive associations, and a more comprehensive replication in an independent sample data set is crucial for the validity of the findings.

## METHODS

### Study participants

The study uses data from 529 participants from the Religious Orders Study (ROS) and 559 participants from the Rush Memory and Aging Project (MAP), two longitudinal cohort studies. ROS and MAP enroll participants free of known dementia. Participants agree to annual clinical evaluations and to donate their brains at death. ROS was initiated in 1994 and enrolls older Catholic priests, nuns, and brothers from nearly 40 groups located in 12 U.S. states (D. A. Bennett et al. 2006; David A. Bennett et al. 2018). By the time the samples were sent for sequencing (end of 2017), 1,437 individuals had completed their baseline assessment. The MAP was established in 1997 and recruits older men and women from retirement communities and individual householders in the greater Chicago area (D. A. Bennett et al. 2006; David A. Bennett et al. 2018, 2012, 2005). As of the end of 2017, 1,967 participants had completed their baseline evaluation.

ROS and MAP are studies conducted by the same team of investigators sharing a common core of measures and procedures, allowing direct comparison between variables and efficient data merging for combined analyses. The follow-up rate for surviving participants surpasses 90%. Both studies were approved by a Rush University Medical Center Institutional Review Board. Each participant provided written informed consent at enrollment and signed the Uniform Anatomical Gift Act.

### Alzheimer’s dementia and cognitive function

Standardized cognitive and clinical assessments are conducted annually by examiners unaware of previous data. The clinical diagnosis for dementia follows the directives provided by the joint working group of the National Institute of Neurological and Communicative Disorders and Stroke and the AD and Related Disorders Association, as described (David A. Bennett, Schneider, Aggarwal, et al. 2006; McKhann et al. 1984). Mild cognitive impairment (MCI) was defined as individuals assessed by the neuropsychologist as cognitively impaired but not diagnosed as having dementia by the examining physician, as previously outlined (D. A. Bennett et al. 2002). Persons without dementia or MCI were designated as having no cognitive impairment (NCI) as described (D. A. Bennett et al. 2002). A final consensus cognitive diagnosis is determined by a neurologist with proficiency in dementia after reviewing select clinical information after death without knowledge of any postmortem data (Julie A. Schneider et al. 2007). For the current association analysis, two binary statuses were used: Alzheimer’s dementia vs no dementia (MCI + NCI) and with cognitive impairment (AD + MCI) vs no cognitive impairment (NCI).

Quantitative measurements of cognitive function were measured yearly. Cognitive evaluations comprise 19 cognitive performance tests that are common to both studies. A Mini-Mental State Examination is employed for descriptive reasons, whereas the Complex Ideational Material from the Boston Diagnostic Aphasia Examination is solely utilized for diagnostic classification. The remaining 17 tests are merged into a comprehensive metric of global cognition. The scores for each test were converted to a composite score (Robert S. Wilson et al. 2015). For the association analysis, measurements were considered proximal to death.

Estimated slopes for global cognition were also included as a measure of cognitive decline. The random slope of global cognition is calculated to the individualized projected pace of change in the global cognition variable across time. This projection is generated through a linear mixed-effects model, with global cognition serving as the longitudinal outcome. The model adjusts for age at baseline, sex, and years of education (De Jager et al. 2012; Oveisgharan et al. 2023). Additionally, as a measure of cognitive resilience, another random slope of global cognition is calculated, controlling for demographics and neuropathologies. The linear mixed-effects model generates the projection using global cognition as the outcome while adjusting for age at baseline, sex, and years of education as demographics, and global AD pathology burden, β-amyloid, PHF tau tangles, gross chronic cerebral infarctions, chronic microinfarctions, Lewy body disease, TDP-43, hippocampal sclerosis, cerebral amyloid angiopathy, cerebral atherosclerosis, and arteriolosclerosis as pathology factors (Boyle et al. 2021).

### Parkinsonism, frailty and motor function

A global Parkinsonian summary score was also tested. The global Parkinsonian summary score is a composite measure of Parkinsonian signs. It is calculated as the average of four separate domains based on a 26-item modified version of the motor portion of the United Parkinson’s Disease Rating Scale (mUPDRS). These domains include bradykinesia, gait, rigidity, and tremor, and they are administered by a trained nurse clinician. These measures are highly reliable and reproducible in both men and women across various cohorts and have been modified to be more applicable to individuals without Parkinson’s disease and easier for non-physicians to administer and score (Aron S. Buchman et al. 2012).

In addition, a total of two motor-related indices were evaluated. A measure of frailty and a composite measure of global motor function. Each of the indices is described below. Frailty is defined as multiple system weaknesses. A continuous composite measure of frailty is based on four components: grip strength, timed walking, body composition (BMI), and fatigue. The raw scores for each component are converted into z-scores using the mean and standard deviation values from all participants at baseline (Aron S. Buchman et al. 2007; A. S. Buchman et al. 2009). The global motor function combines multiple motor tests, including the Purdue Pegboard Test, finger-tapping test, the time and number of steps to cover a distance of 8 feet, the time and number of steps for 360-degree turn, leg and toe stand, grip strength, and pinch strength. Each test’s performance score is converted to a score based on the mean score of all participants at baseline, and then the scores are averaged together to create the composite measure. This measure provides a comprehensive assessment of motor and gait function in individuals (Aron S. Buchman et al. 2009).

### Depression and depressive symptoms

Two measures of depression were evaluated: a binary status of clinical diagnosis of major depressive disorder (MDD) and a quantitative score of depressive symptoms. The clinical diagnosis of major depressive disorder was made by an examining physician at each evaluation. Diagnosis was based on criteria of the Diagnostic and Statistical Manual of Mental Disorders (DSM-III-R), clinical interview with the participant, and a review of responses to questions adapted from the Diagnostic Interview Schedule (David A. Bennett et al. 2004). A binary variable was tested classifying probably or highly probable vs. possible or not present MDD into the presence or absence of MDD. Depressive symptoms were assessed using a modified 10-item version of the Center for Epidemiologic Studies Depression Scale (CES-D) (Kohout et al. 1993; Robert S. Wilson et al. 2002, 2014). Participants were asked whether they experienced each of the ten symptoms frequently in the past week. An overall score was obtained by aggregating the number of symptoms reported.

### Neuropathological evaluations

Systematic assessment of various neurodegenerative and cerebrovascular conditions, including pathological diagnosis of Alzheimer’s disease, Lewy bodies, LATE, hippocampal sclerosis, chronic macroscopic infarcts and microinfarcts, cerebral amyloid angiopathy, atherosclerosis, and arteriolosclerosis were performed as previously reported (Boyle et al. 2019, 2021). The assessments were conducted by examiners who were blinded to all clinical data. A summary of each variable used in the current study is described below:

#### NIA-Reagan diagnosis of AD

The NIA-Reagan diagnosis of Alzheimer’s disease was measured based on a set of consensus recommendations for diagnosing the disease after death, taking in account the presence of both neurofibrillary tangles (Braak) and neuritic plaques (CERAD). This criteria was modified because the neuropathological evaluation is carried out without knowledge of the patient’s clinical information, including a dementia diagnosis. Thus, the level of Alzheimer’s disease pathology is determined by a neuropathologist <u>(Bennett et al. 2006)</u>. In our association analysis, we utilized a dichotomized variable where individuals with an intermediate or high likelihood fulfill the criteria for a pathological diagnosis of Alzheimer’s disease.

#### Burden of neuritic and diffuse plaques and neurofibrillary tangles

Neuric and diffuse plaques were determined through microscopic examination of silver-stained slides from five specific brain regions. The index count in each region is scaled by dividing it by its corresponding standard deviation. The scaled regional measures are then averaged to obtain a summary measure for the plaque burden. The five regions examined are the midfrontal cortex, midtemporal cortex, inferior parietal cortex, entorhinal cortex, and mid-hippocampus CA1. In addition, a global score of AD pathology was also tested as quantitative measure derived from the counts of three silver-stained measures of AD pathologies: neuritic plaques, diffuse plaques, and neurofibrillary tangles. Measures were made in same five regions, and each regional count was scaled by dividing by the corresponding standard deviation. The average of the three measured was used (D. A. Bennett et al. 2003; David A. Bennett, Schneider, Tang, et al. 2006).

#### β-amyloid load

β-amyloid protein is identified by molecularly specific immunohistochemistry and quantified by image analysis in 8 brain regions. The percent area of the cortex occupied by β-amyloid is calculated, and a mean score is determined from 4 or more regions (R. S. Wilson et al. 2007).

#### PHFtau tangle density

Neuronal tangles are identified with phosphorylated Tau protein antibodies (AT8) and quantified in 8 brain regions using systematic sampling by stereology to determine cortical density. A mean tangle score is then calculated from 4 or more regions (R. S. Wilson et al. 2007).

#### TDP-43

TDP-43 cytoplasmic inclusions in neurons and glia are assessed for each of the eight brain regions and scored based on four stages of TDP-43 distribution (ranging from none to involvement of all eight regions). A dichotomized presence or absence is used in these analyses (Nag et al. 2017).

#### Lewy Body disease (LBD)

A pathologic diagnosis of LBD is determined based on four stages of distribution of α-synuclein in the brain. Brain tissue samples from multiple regions are evaluated with α-synuclein immunostaining. The McKeith criteria were modified to assess the presence of LBD in different categories: not present, nigral-predominant, limbic-type, and neocortical-type. A dichotomized version of this variable is used, referring to Lewy bodies present or absent (J. A. Schneider et al. 2012).

#### Arteriolosclerosis

Arteriolosclerosis refers to histological changes observed in small brain vessels during aging, including intimal deterioration, smooth muscle degeneration, and fibrohyalinotic thickening that narrows the vascular lumen. The severity of arteriolosclerosis is evaluated as none, mild, moderate, or severe ( Aron S.Buchman et al. 2011).

#### Cerebral Atherosclerosis

The severity of large vessel cerebral atherosclerosis is visually assessed by examining several arteries and their proximal branches in the circle of Willis. The rating was based on the extent of involvement, including the number of arteries affected and the degree of occlusion. A semiquantitative scale is used: none or possible, mild, moderate, and severe (Arvanitakis et al. 2017).

#### Cerebral Amyloid Angiopathy

A semiquantitative summary of cerebral amyloid angiopathy (CAA) pathology in 4 neocortical regions is calculated using paraffin-embedded sections that were immunostained for β-amyloid using one of three monoclonal anti-human antibodies. Meningeal and parenchymal vessels are assessed for β-amyloid deposition and scored from 0 to 4 based on the extent of circumferential deposition for each region. The CAA score for each region is the maximum of the meningeal and parenchymal CAA scores, which are then averaged across regions to summarize as a continuous measure of CAA pathology (Boyle et al. 2015).

#### Cerebral Infarctions - Gross-Chronic

Neuropathologic evaluations are currently performed at Rush to determine the presence of one or more gross chronic cerebral infarctions. The evaluations are blinded to clinical data and reviewed by a board-certified neuropathologist. The examination documents the age (acute/subacute/chronic), size, and location (side and region) of infarcts visible to the naked eye on fixed slabs. All visible and suspected macroscopic infarcts are dissected for histologic confirmation. A value is one or more gross chronic infarctions vs none (Arvanitakis et al. 2011; J. A. Schneider et al. 2005).

#### Cerebral Infarctions - Micro-Chronic

Neuropathologic evaluations are performed at Rush to determine the presence of one or more chronic microinfarcts, which are chronic microscopic infarctions. The evaluations are blinded to clinical data and reviewed by a board-certified neuropathologist. At least nine regions in one hemisphere are examined for microinfarcts on 6µm paraffin-embedded sections stained with hematoxylin/eosin. The examination includes six cortical regions (midfrontal, middle temporal, entorhinal, hippocampal, inferior parietal, and anterior cingulate cortices), two subcortical regions (anterior basal ganglia, thalamus), and midbrain. Age (acute/subacute/chronic) and location (side and region) of microinfarcts are recorded. A value of 0 indicates no chronic microinfarcts, while a value of 1 indicates the presence of one or more chronic microinfarcts (Arvanitakis et al. 2011).

### WGS data and variant calling

Whole-genome sequencing (WGS) data were previously generated from DNA samples from blood or cortex tissues (De Jager et al. 2018). Briefly, libraries were sequenced on an Illumina HiSeq X sequencer using 2 x 150 bp cycles. Single nucleotide variant and small indel discovery and genotyping were performed utilizing an NYGC automated pipeline, which included alignment to the GRCh37 human reference using the Burrows-Wheeler Aligner and processing using the GATK best-practices workflow. The workflow included marking duplicate reads, local realignment around indels, and using Genome Analysis Toolkit (GATK) base quality score recalibration.

Structural variant discovery and genotyping were also previously described. A combination of seven different software tools, including DELLY (Rausch et al. 2012), LUMPY (Layer et al. 2014), Manta (X. Chen et al. 2016), BreakDancer (K. Chen et al. 2009), CNVnator (Abyzov et al. 2011), BreakSeq (Abyzov et al. 2015), and MELT (Gardner et al. 2017), was applied to identify SVs in each sample. The variants were then merged at the individual level using SURVIVOR (Jeffares et al. 2017) and genotyped at the study level using smoove (GitHub - Brentp/smoove). ROS and MAP were jointly genotyped, resulting in a final set of 72,348 SVs mapped in 1,106 individuals after quality control. Details about the specific SV pipeline steps are described in the original publication (Vialle et al. 2022).

### Linkage disequilibrium with GWAS variants

Linkage disequilibrium between SNPs and SVs was also previously generated (Vialle et al. 2022). A joint call set with 8,566,510 SNPs and 72,348 SVs was used to calculate LD in terms of R2 for all SVs using PLINK and considering a window of 5 Mb.

### Single variant association analysis of SVs and AD/ADRD traits

We performed genome-wide association analysis testing for each common SV with 16 quantitative and eight binary AD/ADRD phenotypes. The variables measuring β-amyloid tangle density, neurofibrillary tangle burden, neuritic and diffuse plaque burden were squared root transformed before the association analysis.

Analysis was performed using SAIGEgds (Scalable and Accurate Implementation of GEneralized mixed model) (Zheng and Davis 2021; Zhou et al. 2018). The method uses the saddlepoint approximation to calibrate the distribution of score test statistics and state-of-the-art optimization strategies to reduce computational costs. SAIGE can analyze large-scale data while controlling for unbalanced case-control ratios and sample relatedness, making it applicable to GWAS for thousands of phenotypes by large biobanks. The method was tested on UK Biobank data and demonstrated its efficiency in analyzing extensive sample data. The SAIGE method involves two main steps: (1) fitting the null logistic mixed model to estimate model parameters using the average information restricted maximum likelihood algorithm, and (2) testing for associations between each genetic variant and phenotype using the saddlepoint approximation method on score test statistics. Several optimization strategies have been applied to make fitting the null logistic mixed model practical for large data sets, such as using the raw genotypes as input and the preconditioned conjugate gradient method to solve linear systems iteratively. These optimizations make SAIGE computationally efficient and applicable to GWAS for thousands of phenotypes by large biobanks.

Here, we performed separate genome-wide scans using 529 participants from ROS and 559 participants from MAP. All tests were controlled by age at death, sex, years of education, five genetic principal components, and the genetic correlation matrix (modeled as a random effect). To combine consistent genetic effects across both studies, a meta-analysis was conducted using METASOFT v.2.0.1 (Han and Eskin 2011). Effect sizes and standard errors of each SV–trait pair were used as input. We carried out a random-effects meta-analysis using the RE2 model, optimized to detect associations under heterogeneity.

### Proteomics data

In our analysis, we utilized ROS/MAP proteomics data to link the impact of SVs with protein abundances. The methods for data generation were previously published in detail (Seifar et al. 2024; Higginbotham et al. 2022; Wingo et al. 2020; Yu et al. 2020). Briefly, frozen DLPFC tissue samples underwent homogenization, followed by the quantification of protein concentrations. Isobaric TMT peptide labels were subsequently introduced and fractioned by high pH. These fractions were then subjected to liquid chromatography-mass spectrometry, and the resulting spectra were cross-referenced with the UniProt database. After quality control to eliminate technical confounders, 10,030 proteins from 971 individuals were available for downstream analysis (Seifar et al. 2024).

## Data availability

All SV site frequency data and complete nominal summary statistics for all phenotypes tested are publicly available on GitHub (https://github.com/RushAlz/ADRD_SV_GWAS). ROS/MAP resources, including individual-level genotyping and phenotypic data can be requested at https://www.radc.rush.edu.

## Code availability

All relevant code used in this study has been provided in a single repository on GitHub (https://github.com/RushAlz/ADRD_SV_GWAS).

## Acknowledgments

We thank the participants of ROS/MAP cohorts for their essential contributions and gift to these projects, as well the investigators and staff at the Rush Alzheimer’s Disease Center. This work has been supported the following National Institute on Aging (NIA) grants: P30AG10161 (DAB), P30AG72975 (JAS), R01AG15819 (DAB), R01AG17917 (DAB), U01AG46152 (PLD, DAB), U01AG61356 (PLD, DAB), R01AG22018 (LLL), U01AG079847 (CG). The funders had no role in study design, data collection and analysis, decision to publish, or preparation of the manuscript.

