## Supplementary Material for "Structural variants linked to Alzheimer’s Disease and other common age-related clinical and neuropathologic traits"

**Supplementary Table 1. Demographic and clinical characteristics of study cohorts**

| | ROS | MAP | ROS/MAP |
|---|---|---|---|
| *n* | 529 | 559 | 1088 |
| Age baseline, mean (SD), y | 78.2 (6.8) | 83.4 (5.8) | 80.9 (6.8) |
| Age at death, mean (SD), y | 88.1 (6.8) | 89.8 (6.0) | 89.0 (6.4) |
| Male sex, No. (%) | 188 (35.5) | 180 (32.2) | 368 (33.8) |
| Educational level, mean (SD), y | 18.1 (3.3) | 14.6 (2.9) | 16.3 (3.6) |
| Length of follow-up, mean (SD), y | 9.1 (5.4) | 5.5 (3.7) | 7.2 (4.9) |
| MMSE score, median (IQR) - at baseline | 29 (27.0-30.0) | 28 (26.0-29.0) | 28 (26.0-29.0) |
| MMSE score, median (IQR) - at death | 24 (14.4-28.0) | 25 (17.0-28.0) | 25 (15.0-28.0) |
| Global cognition score, mean (SD) - at baseline | -0.04 (0.63) | -0.24 (0.76) | -0.14 (0.70) |
| Global cognition score, mean (SD) - at death | -1.00 (1.29) | -0.96 (1.16) | -0.98 (1.23) |
| MCI, No. (%) | 121 (22.9) | 154 (27.5) | 275 (25.3) |
| Dementia, No. (%) | 253 (47.8) | 222 (39.7) | 475 (43.7) |
| NIA-Reagan AD, No. (%)[a] | 329 (62.2) | 361 (64.6) | 690 (63.4) |
| Global AD pathologic score, median (IQR) | 0.67 (0.18-1.18) | 0.65 (0.19-1.20) | 0.67 (0.19-1.20) |
| Neuritic plaques score, median (IQR) | 0.69 (0.03-1.31) | 0.71 (0.06-1.35) | 0.71 (0.05-1.34) |
| Diffuse plaques score, median (IQR) | 0.66 (0.11-1.31) | 0.54 (0.07-1.11) | 0.60 (0.08-1.19) |
| Neurofibrillary tangles score, median (IQR) | 0.38 (0.13-0.84) | 0.39 (0.15-0.93) | 0.39 (0.14-0.89) |
| Macroscopic infarcts, No. (%) | 188 (35.5) | 202 (36.1) | 390 (35.8) |
| Microinfarcts, No. (%) | 145 (27.4) | 162 (29.0) | 307 (28.2) |
| Neocortical Lewy bodies, No. (%) | 66 (12.9) | 70 (13.0) | 136 (12.9) |
| TDP-43, No. (%)[b] | 155 (33.6) | 187 (34.8) | 342 (34.3) |
| Hippocampal sclerosis, No. (%) | 39 (7.5) | 53 (9.5) | 92 (8.5) |
| Amyloid angiopathy, No. (%)[c] | 183 (36.3) | 190 (34.2) | 373 (35.2) |
| Atherosclerosis, No. (%)[c] | 204 (39.0) | 188 (33.7) | 392 (36.3) |
| Arteriolosclerosis, No. (%)[c] | 161 (30.8) | 212 (38.0) | 373 (34.5) |

Abbreviations: AD, Alzheimer's disease; IQR, interquartile range; MCI, mild cognitive impairment; MMSE, Mini-Mental State Examination; NIA, National Institute on Aging; TDP-43, transactive response DNA-binding protein 43.

[a] Intermediate or high likelihood.

[b] Inclusion beyond the amygdala.

[c] Moderate or severe.

**Supplementary Table 2. Variables and categories of each phenotype tested**

| variable | family | category | description |
|---|---|---|---|
| cogn_global_lv | Continuous | Cognition | Global cognitive function |
| cogng_demog_slope | Continuous | Cognition | Cognitive decline |
| cogng_path_slope | Continuous | Cognition | Cognitive resilience |
| tangles_sqrt | Continuous | Pathology | Tangle density |
| nft_sqrt | Continuous | Pathology | Neurofibrillary tangle |
| amyloid_sqrt | Continuous | Pathology | Beta-Amyloid |
| plaq_n_sqrt | Continuous | Pathology | Neuritic plaques |
| plaq_d_sqrt | Continuous | Pathology | Diffuse plaques |
| tdp_43_binary | Binary | Pathology | TDP-43 |
| ci_num2_gct | Binary | Vascular Pathology | Cerebral Infarctions (Gross) |
| ci_num2_mct | Binary | Vascular Pathology | Cerebral Infarctions (Micro) |
| arteriol_scler | Continuous | Vascular Pathology | Arteriolosclerosis |
| caa_4gp | Continuous | Vascular Pathology | Cerebral amyloid angiopathy |
| cvda_4gp2 | Continuous | Vascular Pathology | Cerebral Atherosclerosis |
| ad_dementia_status | Binary | Cognition | Alzheimer's Dementia |
| cog_impairment_status | Binary | Cognition | Mild cognitive impairment |
| ad_reagan | Binary | Pathology | NIA-Reagan diagnosis of AD |
| gpath | Continuous | Pathology | Global AD pathology |
| parksc_lv | Continuous | Motor | Global parkinsonian score |
| dlbany | Binary | Pathology | Lewy Body disease |
| r_depres_status | Binary | Depression | Major Depressive Disorder |
| cesdsum_lv | Continuous | Depression | Depressive symptoms (CES-D) |
| frailty_z_lv | Continuous | Motor | Frailty |
| motor10_lv | Continuous | Motor | Motor functions |

**Supplementary Table 3. AD GWAS loci in LD with SVs associated with ADRD traits (*P* < 0.05)**

| AD GWAS locus | SV id | SV info | SV LD with lead SNP (R2) | Best phenotype associated with SV | P-value of SV-phenotype association |
|---|---|---|---|---|---|
| chr6:32075406-33083813 (HLA-DRB5–) | DEL_25745 | DEL chr6:32512662 (len:-405 maf:0.310688) | 0.91 | Tangle density | 3.74E-02 |
| chr6:32075406-33083813 (HLA-DRB5–) | DEL_25747 | DEL chr6:32514898 (len:-1399 maf:0.312047) | 0.91 | Tangle density | 3.04E-02 |
| chr6:32075406-33083813 (HLA-DRB5–) | DEL_25752 | DEL chr6:32523647 (len:-6180 maf:0.468661) | 0.59 | Major Depressive Disorder | 7.16E-02 |
| chr6:32075406-33083813 (HLA-DRB5–) | DEL_25737 | DEL chr6:32492661 (len:-60144 maf:0.449367) | 0.55 | Major Depressive Disorder | 3.64E-02 |
| chr6:32075406-33083813 (HLA-DRB5–) | DEL_25768 | DEL chr6:32624892 (len:-950 maf:0.433544) | 0.52 | Cognitive decline | 6.77E-02 |
| chr6:32075406-33083813 (HLA-DRB5–) | DEL_25739 | DEL chr6:32494986 (len:-336 maf:0.32361) | 0.51 | Cognitive decline | 5.16E-02 |
| chr6:32075406-33083813 (HLA-DRB5–) | DEL_25758 | DEL chr6:32561955 (len:-2671 maf:0.175407) | 0.51 | Tangle density | 6.39E-02 |
| chr6:32075406-33083813 (HLA-DRB5–) | DEL_25759 | DEL chr6:32571348 (len:-319 maf:0.44349) | 0.50 | Major Depressive Disorder | 3.19E-02 |
| chr6:32075406-33083813 (HLA-DRB5–) | DUP_4026 | DUP chr6:32490722 (len:35810 maf:0.424051) | 0.40 | Tangle density | 1.01E-02 |
| chr6:32075406-33083813 (HLA-DRB5–) | DUP_4027 | DUP chr6:32494883 (len:43223 maf:0.429928) | 0.39 | Major Depressive Disorder | 3.28E-03 |
| chr6:32075406-33083813 (HLA-DRB5–) | DEL_25746 | DEL chr6:32514825 (len:-26282 maf:0.19575) | 0.35 | Major Depressive Disorder | 2.44E-02 |
| chr6:32075406-33083813 (HLA-DRB5–) | ALU_3734 | ALU chr6:32589834 (len:281 maf:0.116637) | 0.35 | Major Depressive Disorder | 1.24E-01 |
| chr6:32075406-33083813 (HLA-DRB5–) | DEL_25761 | DEL chr6:32592405 (len:-127 maf:0.214932) | 0.35 | Tangle density | 2.65E-02 |
| chr6:32075406-33083813 (HLA-DRB5–) | DEL_25765 | DEL chr6:32622513 (len:-337 maf:0.438065) | 0.32 | Cerebral Infarctions (Micro) | 4.03E-02 |
| chr6:32075406-33083813 (HLA-DRB5–) | DEL_25763 | DEL chr6:32614951 (len:-401 maf:0.438065) | 0.32 | Major Depressive Disorder | 1.14E-02 |
| chr6:32075406-33083813 (HLA-DRB5–) | DEL_25767 | DEL chr6:32623524 (len:-1077 maf:0.426763) | 0.31 | Cognitive resilience | 4.49E-02 |
| chr6:32075406-33083813 (HLA-DRB5–) | SVA_296 | SVA chr6:32514612 (len:1267 maf:0.11786) | 0.30 | Arteriolosclerosis | 9.51E-02 |
| chr6:32075406-33083813 (HLA-DRB5–) | DEL_25764 | DEL chr6:32615970 (len:-4223 maf:0.304702) | 0.29 | Beta-Amyloid | 1.42E-01 |
| chr6:32075406-33083813 (HLA-DRB5–) | DEL_25705 | DEL chr6:32450599 (len:-86768 maf:0.310127) | 0.28 | Cognitive resilience | 6.13E-03 |

| | | | | | |
|---|---|---|---|---|---|
| chr6:32075406-33083813 (HLA-DRB5–) | INS_5722 | INS chr6:32664240 (len:57 maf:0.113472) | 0.28 | Frailty | 7.59E-02 |
| chr6:32075406-33083813 (HLA-DRB5–) | ALU_3719 | ALU chr6:32449295 (len:280 maf:0.327758) | 0.27 | Motor functions | 5.01E-02 |
| chr6:32075406-33083813 (HLA-DRB5–) | DEL_25748 | DEL chr6:32516620 (len:-24772 maf:0.242315) | 0.27 | Tangle density | 2.91E-02 |
| chr6:32075406-33083813 (HLA-DRB5–) | DUP_4025 | DUP chr6:32488856 (len:35679 maf:0.26085) | 0.25 | Neurofibrillary tangle | 1.20E-01 |
| chr6:32075406-33083813 (HLA-DRB5–) | DEL_25716 | DEL chr6:32455073 (len:-23356 maf:0.279292) | 0.23 | Frailty | 1.42E-01 |
| chr6:32075406-33083813 (HLA-DRB5–) | DEL_25695 | DEL chr6:32416934 (len:-105 maf:0.427667) | 0.23 | Major Depressive Disorder | 1.69E-02 |
| chr6:46931284-48052180 (CD2AP) | DEL_25938 | DEL chr6:47564729 (len:-2187 maf:0.0691682) | 0.22 | Mild cognitive impairment | 8.86E-02 |
| chr7:11768758-12769593 (TMEM106B) | DEL_27662 | DEL chr7:12281711 (len:-343 maf:0.424955) | 0.96 | Tangle density | 7.72E-04 |
| chr10:123672912-124672912 (PLEKHA1) | DEL_3676 | DEL chr10:124216821 (len:-461 maf:0.230561) | 0.23 | Global parkinsonian score | 1.16E-02 |
| chr11:46880340-48057871 (CELF1) | DEL_4449 | DEL chr11:47887667 (len:-535 maf:0.443942) | 0.56 | Cerebral Infarctions (Micro) | 1.19E-02 |
| chr11:46880340-48057871 (CELF1) | ALU_6357 | ALU chr11:47806655 (len:280 maf:0.347197) | 0.39 | Cerebral Atherosclerosis | 6.04E-02 |
| chr11:46880340-48057871 (CELF1) | DEL_4444 | DEL chr11:47563238 (len:-96 maf:0.368897) | 0.39 | Mild cognitive impairment | 4.43E-02 |
| chr12:113219788-114219788 (TPCN1) | INS_11268 | INS chr12:113724222 (len:69 maf:0.0384268) | 0.67 | Lewy Body disease | 4.35E-02 |
| chr14:52798853-53900629 (FERMT2) | INV_1034 | INV chr14:53274515 (len:179 maf:0.084991) | 0.41 | Cerebral Atherosclerosis | 1.28E-02 |
| chr15:58522615-59557023 (ADAM10) | DEL_9672 | DEL chr15:58912865 (len:-329 maf:0.344033) | 0.39 | Global parkinsonian score | 1.31E-01 |
| chr16:19308163-20308163 (IQCK) | DEL_10481 | DEL chr16:19945550 (len:-22029 maf:0.142405) | 0.44 | Major Depressive Disorder | 2.34E-03 |
| chr17:17559454-18559454 (MYO15A) | DEL_11664 | DEL chr17:18270187 (len:-1505 maf:0.0863472) | 0.37 | TDP-43 | 7.02E-03 |

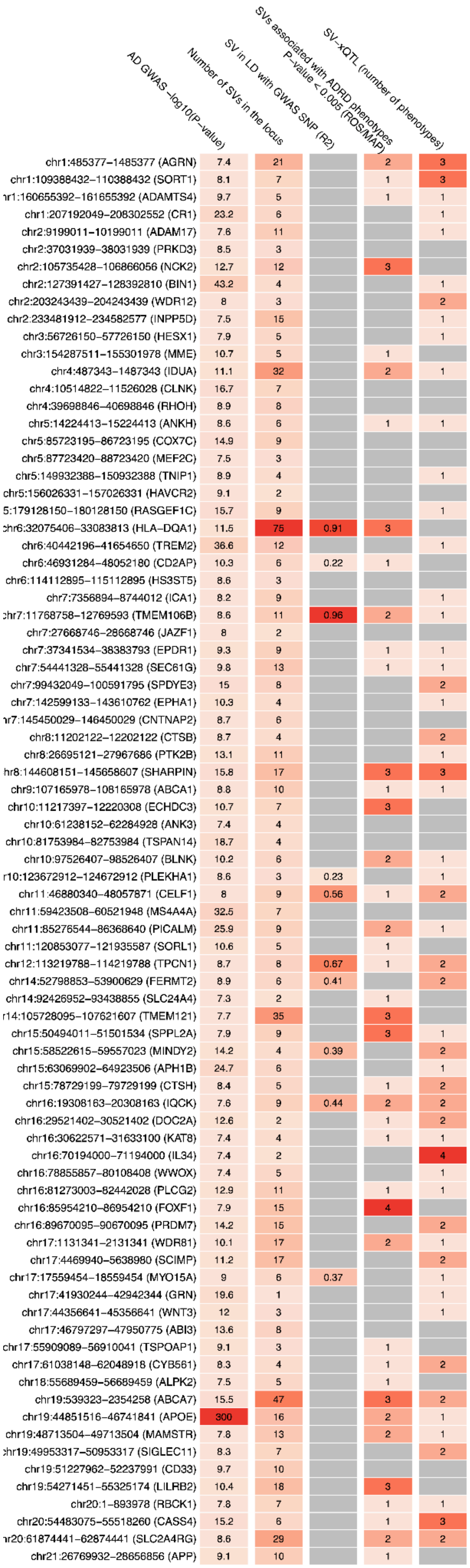


**Supplementary Figure S1. Overview of SVs in 81 AD GWAS loci.** Heatmap shows for each of the AD loci (row): 1) the strength of the association in the GWAS study; 2) the number of SVs in the region; 3) the strength of LD between the lead GWAS SNP and an SV (highest R2); 4) the number of ADRD phenotypes associated in the SV-GWAS tests; 5) the number of molecular phenotypes linked to an SV found in each locus (SV-xQTL).
